# Meta-Analysis of Clinical Phenotype and Patient Survival in Neurodevelopmental Disorder with Microcephaly, Arthrogryposis, and Structural Brain Anomalies Due to Bi-allelic Loss of Function Variants in SMPD4

**DOI:** 10.1101/2022.10.08.22280875

**Authors:** Dean Marchiori

**Author notes:** Corresponding author: First Author^1^.

## Abstract

A recently described, rare genetic condition known as Neurodevelopmental Disorder with Microcephaly, Arthrogryposis, and Structural Brain Anomalies (NEDMABA) has been identified in children with bi-allelic loss-of-function variants in *SMPD4*. The progression of this condition is not well understood with the limited case reports described so far exhibiting a severe and clinically diverse phenotype. A gap exists in the understanding of associations present in the heterogenous features of the clinical phenotype, and the expected survival probabilities of affected individuals. This is driven in part to the paucity of analysis-ready data on reported cases. This analysis aims to collate and standardise available case reports into a common dataset, to analyse and identify meaningful clusters in the clinical phenotype, and to quantify the survival probability for children with NEDMABA. To overcome the challenge of multidimensional data on very few subjects, we employ Multiple Correspondence Analysis (MCA) as a dimension reduction technique, which is then subject to cluster analysis and interpretation. To account for censoring in the data, Kaplan-Meier estimation is formulated to calculate patient survival time. The analysis correctly detected the classic phenotype for this condition, as well as a new distinct feature-cluster relating to findings of vocal cord paralysis, feeding dysfunction and respiratory failure. The survival probability for those affected was found to decline sharply in early infancy with median survival of 150 days, but with some surviving as long as 12.5 years. This wide range of outcomes is provisionally associated with different variant types however this conclusion could not be validated based on very low sample sizes. An `R` package called `SMPD4` was developed to publish standardised analysis-ready datasets used in this study. This analysis represents the first of its kind to help describe associations and trajectories of individuals with this newly reported condition, despite challenges with sparse and inconsistent data. This analysis can provide clinicians and genetic counsellors with better information to aide in decision making and support for families with this rare condition.

## INTRODUCTION

Neurodevelopment disorders are a diverse and heterogeneous group of conditions impacting the development of the nervous system, brain function, physical development and learning ability. A recently identified condition known as Neurodevelopmental Disorder with Microcephaly, Arthrogryposis, and Structural Brain Anomalies (NEDMABA) ([MIM:618622](https://omim.org/entry/618622)) has been described in children with bi-allelic loss-of-function variants in *SMPD4* Magini et al. (2019). Sphingomyelinases such as *SMPD4* play an important cellular role by hydrolyzing sphingomyelin into ceramide and phosphorylcholine. *SMPD4* specifically encodes one of 4 neutral sphinogmyelinases, nSMase3 (MIM: 610457). So far, less than 50 cases of this rare disorder have been reported in the literature. Clinical phenotype data of these cases is largely heterogeneous with severe neurological complications and early demise a key feature. This makes the identification, diagnosis and ongoing management of cases a challenge for patients, their families and medical practitioners.

A detailed study by Magini et al. (2019) involved 12 unrelated families with 32 individuals (21 with detailed clinical information). Presentations were of microcephaly, simplified gyral pattern of the cortex, hypomyelination, cerebellar hypoplasia, congenital arthrogryposis, and early fetal/postnatal demise. Despite this being the largest cohort studied, the clinical features and survival times among participants varied greatly. Three missense changes were noted in the study with affected children in these families often showing a milder presentation suggestive of possible residual function. In these cases individuals were able to develop independent motor skills, have mild intellectual disability and arthrogryposis without evidence of simplified gyral patterns on brain MRI. Other patients with truncating variants are shown to have more severe presentations, while a range of additional significant clinical features were reported involving dysmorphic facial features, seizure, vocal cord paralysis and hearing impairment. Ravenscroft et al. (2021) studied a family from Melbourne exhibiting features involving arthrogryposis multiplex congenita, complex brain malformations, small for gestation age and hypoplasia of the corpus callosum. In two of the three related cases were additional features of microcephaly, congenital encephalopathy, cerebellar malformation and hypoplasia and hypomyelination. A case study from China is described in Ji et al. (2022) involving a girl presenting in infancy with intrauterine growth restriction, microcephaly, postnatal developmental delay, arthrogryposis, hypertonicity, seizure, and hypomyelination on brain magnetic resonance imaging. The authors report parallels with two cases showing the same homozygous null variant. An individual reported in Monies et al. (2019) presented with distinct symptoms of brain atrophy and skeletal dysplasia whereas the case in Magini et al. (2019) with the same variant exhibited more typical clinical features. A recent study by Bijarnia-Mahay et al. (2022) presents the case of a 22-month old girl presenting with the typical phenotype of neurodevelopmental delay, prenatal onset growth failure, arthrogryposis, microcephaly and brain anomalies including severe hypomyelination, simplified gyral pattern and hypoplasia of corpus callosum and brainstem. Notably, there was also additional non-typical clinical findings of nystagmus and visual impairment secondary to macular dystrophy and retinal pigment epithelial stippling at posterior pole.

Further work to collate and analyse data relating to NEDMABA is challenging. While Magini et al. (2019) included a supplementary clinical phenotype data set in their study, these data are not suitable for statistical analysis. Many of the clinical features are represented as text descriptions and a variety of non-standard terminologies are applied between cases, making machine interpretation of the data difficult. Other studies (Ji et al., 2022; Bijarnia-Mahay et al., 2022) present only written case reports with some tabulated summaries. While larger studies (Ravenscroft et al., 2021; Monies et al., 2019) focus more on genetic data, with less detail included on the clinical presentation of individuals. To overcome these data challenges, Wickham (2014) proposes a data structure know as ‘tidy’ data which organises each observation in a row, with each feature as its own column and each value in just one cell. In this case detailed text-based data can be transformed to high dimensional binary indicators of key clinical features. This structure is conducive to effective statistical analysis and integrates deliberately with the *tidyverse* (Wickham, 2014) collection of packages for data analysis in R (R Core Team, 2022).

Further analysis to better describe rare genetic conditions was explored in Díaz-Santiago et al. (2020) in the context of large scale genotype-phenotype analysis. The authors argue patients with rare disorders (often in small samples) frequently present with varied symptoms that do not match exactly with the described phenotype. When considering the situation of low sample size data with many features, methods in the field of multivariate statistical analysis are commonly deployed. Here the aim is to find substructure in the data, or a simple representation of the multi-dimensional space. Methods, such as Multiple Correspondence Analysis (Le Roux and Rouanet, 2010) are appropriate for high dimensional data comprised of binary indicators. This technique is cited in many studies in the application of dimension reduction and clustering of comorbidities and phenotypes from disease (Han et al., 2018; Costa et al., 2013).

In terms of patient survival, methods in statistical survival analysis are well suited and commonly used in the field of genomics Chen et al. (2014). These methods in particular provide a mechanism for dealing with right-censoring, a common feature of clinical studies where the clinical outcome of interest may not be known by the end of the study period.

An open research question exists around the clinical pathway and survival time of children exhibiting bi-allelic loss-of-function variants in *SMPD4*. With current research highlighting a diverse and heterogeneous phenotype, the relationship between these diverse features is not well understood. Furthermore, the survival time of affected children has not been extensively analysed despite early-demise featuring as a severe outcome. While a connection has been highlighted between some missense variants and a milder presentation with longer survival (Magini et al., 2019), this hypothesis has not been analysed further.

This analysis has three key aims. First, to collate and transform the variety of early case reports and studies on clinical phenotype data for this novel variant into an analysis-ready *tidy* data set. Secondly, to conduct analysis on the associations between commonly reported clinical features. Finally, to statistically quantify the expected survival time of children with NEDMABA based on the current case reports.

This will be the first in-depth analysis of this novel variant, which aims to produce statistical findings to better understand this newly described condition. This research will assist clinicians understand the various presentations of this challenging new condition. In addition, genetic counselling of families with affected children will benefit from enhanced analysis on outcomes of existing reported cases.

## METHODS

### SMPD4 Data Package

The data contained in Magini et al. (2019) “Summary of SMPD4-Related Clinical Phenotype” is an excel spreadsheet tabulating each of the 21 individuals from the study with clinical details recorded. The file has 21 columns (one for each individual), and 64 rows (one for each phenotype or clinical remark).

The data were read into `R` (R Core Team, 2022) without any changes, so as to preserve the reproducibility of the data transformation steps. The data were transposed to ensure it could be presented as *tidy* formatted data (Wickham, 2014). This requires that each variable (clinical phenotype) forms a column; each observation (individual) forms a row and each type of observational unit forms a table (every cell has just one item). This resulted in a long-formatted data set of 21 rows (individuals) and 64 columns (clinical features). In many cases, key clinical information was entered as free-text descriptions, which rendered any attempt of meaningful analysis impractical (Table 1). In these cases, the text was tokenised by separating the list of clinical observations at each comma and forming a binary indicator column noting its presence ‘1’ or absence ‘0’ (Table 2).

**Table 1.** Example of non-tidy free-text descriptions of features.

| Family 1- Individual 1 | Family 1- Individual 4 |
| --- | --- |
| short palpebral fissures, large ears, simple helices | short palpebral fissures, receding forehead |

**Table 2.** Example of tidy formatted data where individuals are transposed into rows and text into binary indicators

| id | short palpebral fissures | large ears | simple helices | receding forehead |
| --- | --- | --- | --- | --- |
| Family 1- Individual 1 | 1 | 1 | 1 | 0 |
| Family 1- Individual 4 | 1 | 0 | 0 | 1 |

In the case where synonymous terms are present, these indicator columns are consolidated and merged into one indicator column to prevent duplication e.g. {bilateral cleft lips, bilateral cleft lip, cleft lip b l} -*>* {bilateral cleft lip}.

Data type conversion and categorical level standardisation was performed to ensure the data were in consistent and appropriate data types. For example, ‘Gender’ was not consistently coded, and ‘Birth Weight’ was encoded as a text string rather than a more useful numeric format. (Table 3).

**Table 3.** Example of inconsistent coding or sub-optimal data types

| Gender | Birth Weight |
| --- | --- |
| male | 2175 grams (- 2.5 SD) |
| female | 2045 g (-3 SD) |
| Female | 2300 gram (-2 SD) |
| Female fetus | n.a. |

This format was used as a template to align other written case reports identified in the literature (Ravenscroft et al., 2021; Monies et al., 2019; Bijarnia-Mahay et al., 2022; Ji et al., 2022). These were manually entered to conform to this template to allow the data to be combined for further analysis.

The final data set consisted of 28 observations (one per individual) and 152 variables (one per clinical feature). These variables were comprised of qualitative text descriptions, binary indicators of features as described above and other fields including background information on the patient and various bio markers. A full list of this data set is available in Table 11. These data sets were compiled into a publicly available `R` Package called `SMPD4` (Marchiori, 2022) to allow for reproducibility and sharing.

### Multiple Correspondence

The data on subjects above is subset to include only variables that can be transformed into binary indicator variables representing the presence or absence of a given clinical feature. Only those features that appeared in more than one case report were included to minimise the influence of isolated features.

Dimension reduction on this wide data set was performed using Multiple Correspondence Analysis (MCA) (an analogy to Principle Component Analysis (PCA) for categorical data) (Le Roux and Rouanet, 2010) in order to arrive at a set of meaningfully small dimensions that account for most of the variation in the data. This computation was carried out using the *FactoMineR* R package (Lê et al., 2008).

We let these data be **X** which is comprised of a set of individuals *I* and a set of features *Q* such that the *q*_*th*_ feature has *K*_*q*_ levels. The sum of all categories 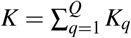 defines the dimensionality of **X** as an *I × K* matrix. This resulted in a data set *X*_28*×*61_ indicator matrix of 61 clinical features across 28 individuals. Taking *δ*_*ik*_ = 1 if subject *i* has feature *k* and *δ*_*ik*_ = 0 if the subject does not, we are left with the completely disjunctive table **X** = *I ×K* of {0, 1}. Letting the sum of all entries of **X** be *N*, we have **Z** = *N*^−1^**X**. We can introduce two diagonal matrices **D**_**r**_ = *diag*(**r**) and **D**_**c**_ = *diag*(**c**) where **r** and **c** are the vectors of row sums and column sums of **Z** respectively.

Computing MCA involves taking the Singular Value Decomposition:

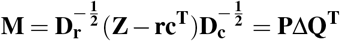

where ∆ are the singular values and Λ = ∆^2^ is the matrix of eigenvalues.

This results in the row and column factor scores respectively as:

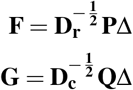

The *contribution* of each category was computed for each of the principal axis per Le Roux and Rouanet (2010). This metric identifies the proportion of variance of that axis due to the point and is defined as:

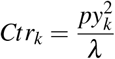

Where *p* represents the point weighting, *y* is the coordinate relative to the principal axes of variance *λ* .

The coordinates on the first few principal axes were inspected for any outliers or unusual data before being graphically interpreted and subject to further analysis.

### Cluster Analysis

The MCA coordinates for the first 4 principal dimensions were subject to clustering in order to find groups of features that were internally homogeneous and externally heterogeneous. This was computed in `R` using the `clValid` package (Brock et al., 2008).

A range of clustering algorithms were compared including AGNES hierarchical agglomerative (Kaufman and Rousseeuw, 2009), k-means (Hartigan and Wong, 1979) and self-organising maps (SOM) (Kohonen, 2012). A sensible range of clusters from 2-5 were trialed for each clustering algorithm. The results for each method were evaluated using three measures of stability: Average proportion of non-overlap (APN), Average Distance (AD) and Average Distance Between Means (ADM) (Datta and Datta, 2003). Stability measures compare the full clustering result with a result based on dropping each of the columns one at a time. These metrics were selected to represent each data point (clinical feature) with a number of MCA dimensions. A good cluster solution is one that is robust and meaningful across all of these dimensions (Figure 5). The optimal method and cluster number was selected based on a trade off between the best performing method using the evaluation metrics and a sensible partitioning of the data for this analysis (Table 5).

**Table 4.** Example of consistent and appropriate coding or data types

| Gender | Birth Weight (g) |
| --- | --- |
| male | 2175 |
| female | 2045 |
| female | 2300 |
| female | NA |

**Table 5.**
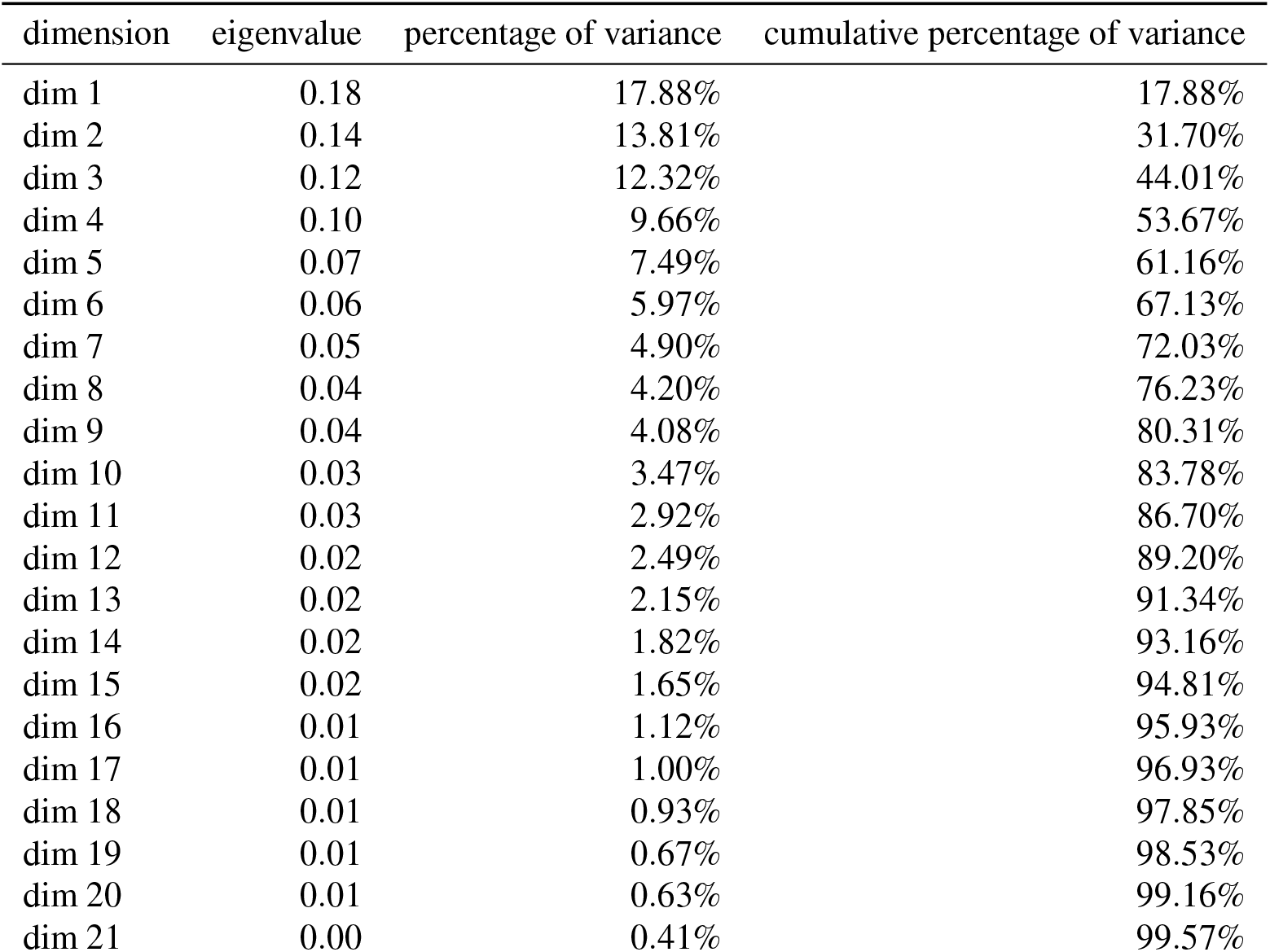
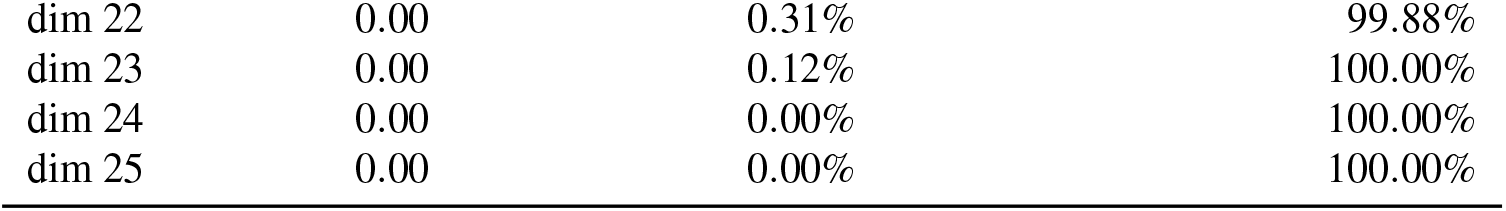
Eigenvalues and percentage of variance explained by the MCA principal axes

### Survival Analysis

To model survival probability for all subjects in the combined data, the data were subsetted to include *survival time* which is either the number of days the individual survived for, or the age in days at last follow up, and *deceased* a numeric event indicator which is equal to 1 if the subject is deceased and 0 if the subject was still alive at the time the respective study had concluded. The Kaplan-Meier estimator (Kaplan and Meier, 1958) was used to estimate the survival function of the data.

The estimator is calculated as:

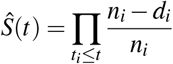

where *t*_*i*_ is some event time, *d*_*i*_ represents the number of events (here deceased subjects) and *n*_*i*_ indicating the individuals known to have survived up to *t*_*i*_The baseline estimator of all individuals was calculated using the *survival* package in `R` (Terry M. Therneau and Patricia M. Grambsch, 2000). Next the Kaplan-Meier estimator was stratified by the type of genetic variation reported in the literature. A log-rank test was performed to detect differences in the survival curves using methods from Harrington and Fleming (1982), again implemented in the *survival* `R` package.

## RESULTS

### MCA

The initial results of the MCA were inspected with a bi-plot and identified two subjects (17, 18) from the data that were clear outliers and sat significantly beyond the 95% confidence ellipse (Figure 1). These individuals represented two twins from Magini et al. (2019) who were described with a much milder phenotype and were subject to a missense variation in *SMPD4*. Further analysis of this influence is conducted below. For now, these subjects were removed to allow for more meaningful analysis of other features.

**Figure 1.**
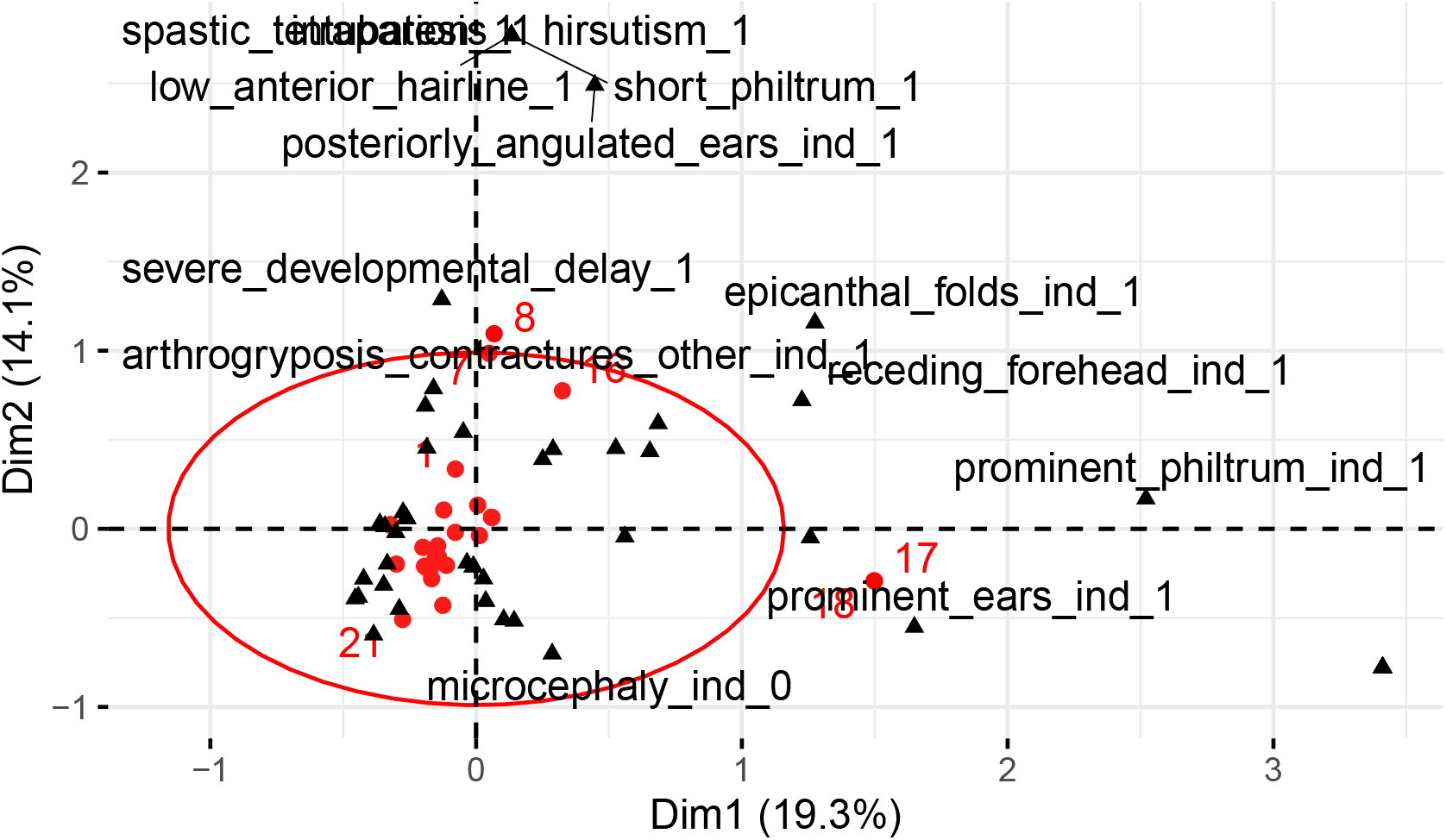
A bi-plot of the first two principal axes with grey circles indicating the individual’s projected position and black triangles indicating the variables. A 95% confidence ellipse is plotted in red around the origin.

Repeating MCA for the remaining subjects shows the first dimension (*λ*_1_ = 0.18) accounts for 17.88% of the variance in the data with the first 4 dimensions of the MCA analysis accounting for 54% of the variance (Table 5).

The *contribution* of the categories were ranked within each of the first four principal dimensions in Figure 2. The first dimension is dominated by isolated cases with highly specific facial dysmorphisms such as epicanthal folds, short philtrum and low anterior hairline. The second MCA dimension is categorised by feeding and respiratory dysfunction with vocal cord palsy, tracheostomy, tube feeding and GERD. Examining the bi-plot of the first two principal axes (Figure 3) illustrates the contrasting sources of variation between conditions relating to airway and swallowing function and predominantly facial features and structural brain anomalies such as as cerebellar hypoplasia, development delay. The third dimension is characterised by atypical, complex features such as patent ductus arteriosus, respiratory infections, small for gestational age and further facial features. These are contrasted by the fourth principal dimension which is strongly categorised by respiratory distress including stridor and hypercapnia in the absence of typical presentations of simplified gyral pattern, intrauterine growth retardation and microcephaly.

**Figure 2.**
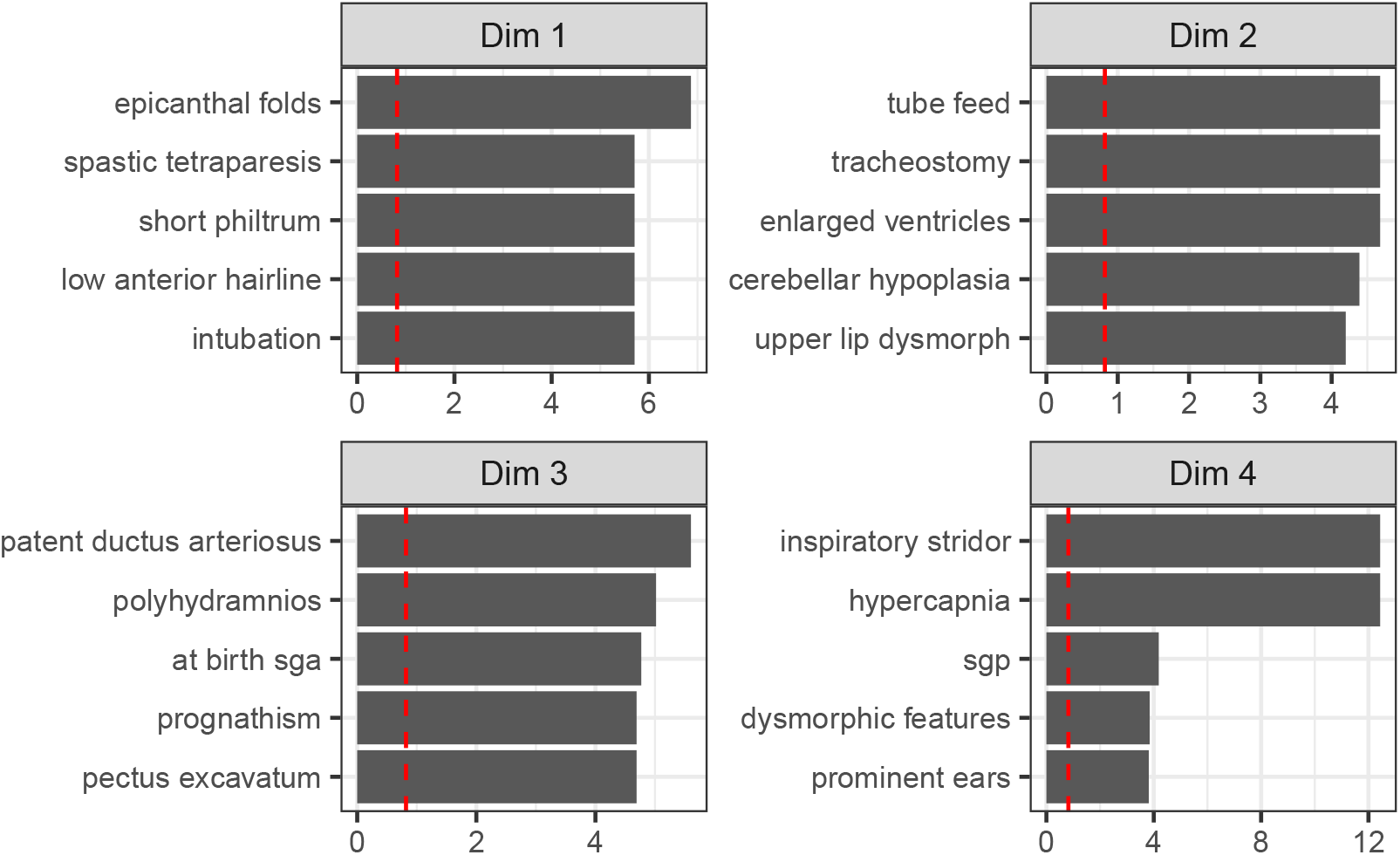
Top 5 feature categories from MCA analysis for the first four MCA dimensions. The red dashed line indicates the mean contribution value.

**Figure 3.**
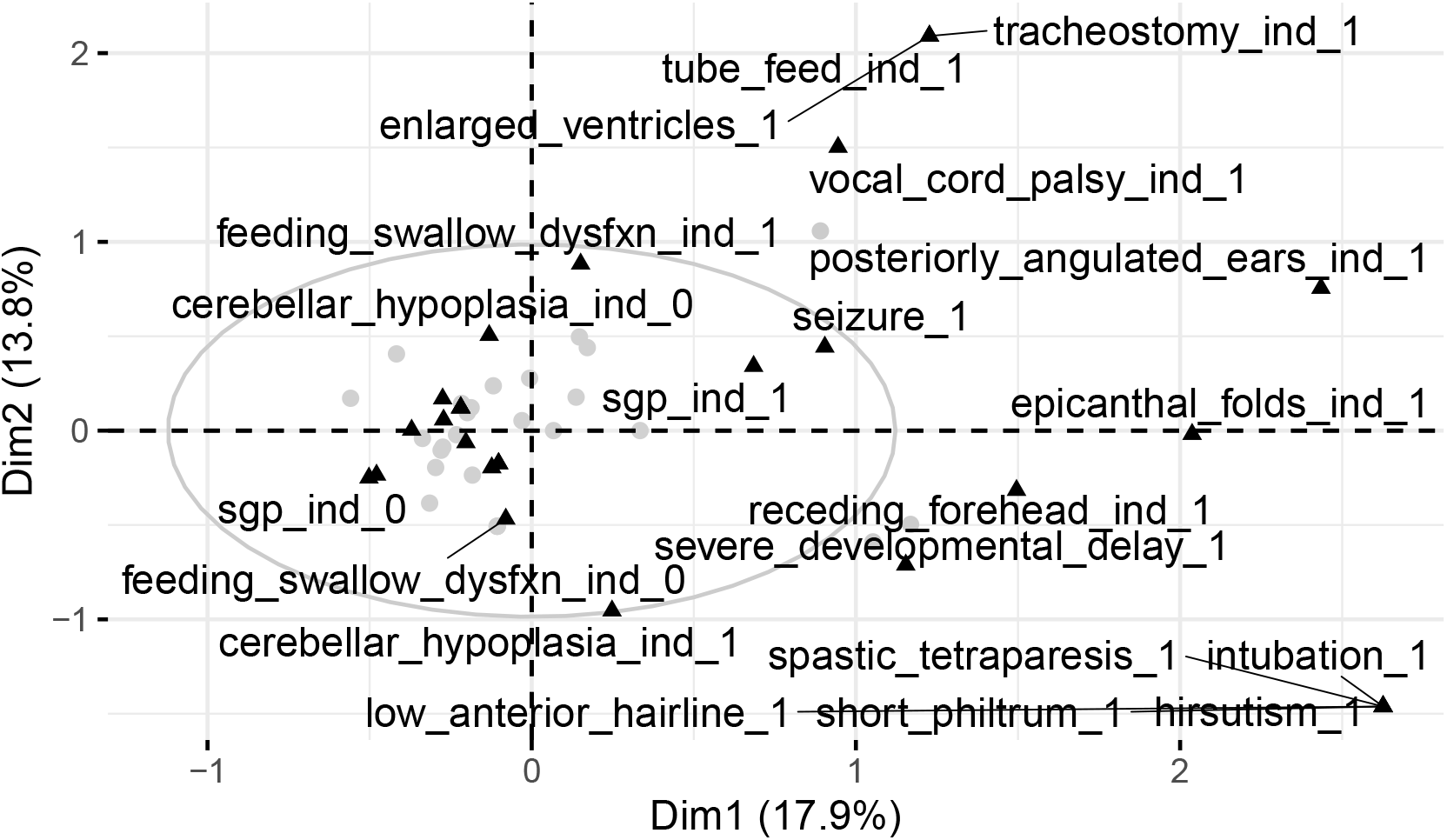
A bi-plot of the first two principal axes with grey circles indicating the individual’s projected position and black triangles indicating the variables. A 95% confidence ellipse is plotted in grey around the origin.

### 0.1 Cluster Analysis

Cluster validation measures resulted in either 2 or 5 clusters as the optimal choice (Table 6). In order to partition the data into meaningful groupings, k-means with 5 clusters was selected. This reflects the optimal choice for both AD and FOM cluster metrics. The results are summarised in Table 7 and a scatter plot of the cluster results is shown in Figure 6 is projected on the first two principal dimensions. This highlights a core cluster of classic features of NEDMABA such as microcephaly, hypomyelination, arthrogryposis and feet deformity and structural brain anomalies. A separate and adjacent cluster is formed of airway and feeding related conditions involving vocal cord paralysis, tracheostomy, tube feeding and enlarged ventricles. Three other smaller satellite clusters exists where highly distinctive features were associated with a single individual or family. A full articulation of feature to cluster assignment is given in Table 9.

**Table 6.** Optimal cluster numbers and methods based on a range of stability validation metrics.

| Metric | Score | Method | Clusters |
| --- | --- | --- | --- |
| APN | 0.07 | agnes | 2 |
| AD | 1.38 | kmeans | 5 |
| ADM | 0.33 | kmeans | 2 |
| FOM | 0.65 | kmeans | 5 |

**Table 7.** A summary table to cluster membership showing the cluster number and number of features in each cluster and a sample of features that have membership in each.

| Cluster No. | No. Features | Sample Features in Cluster<sup>1</sup> |
| --- | --- | --- |
| 1 | 7 | polyhydramnios, short palpebral fissures, patent ductus arteriosus, thin lips |
| 2 | 11 | vocal cord palsy, respiratory distress, tracheostomy, posteriorly angulated ears |
| 3 | 7 | epicanthal folds, receding forehead, spastic tetraparesis, intubation |
| 4 | 21 | microcephaly, sgp, iugr, talipes |
| 5 | 6 | hypercapnia, inspiratory stridor, oligohydramnios, prominent ears |

<sup>1</sup>A sample of 4 features shown to illustrate cluster membership

**Figure 4.**
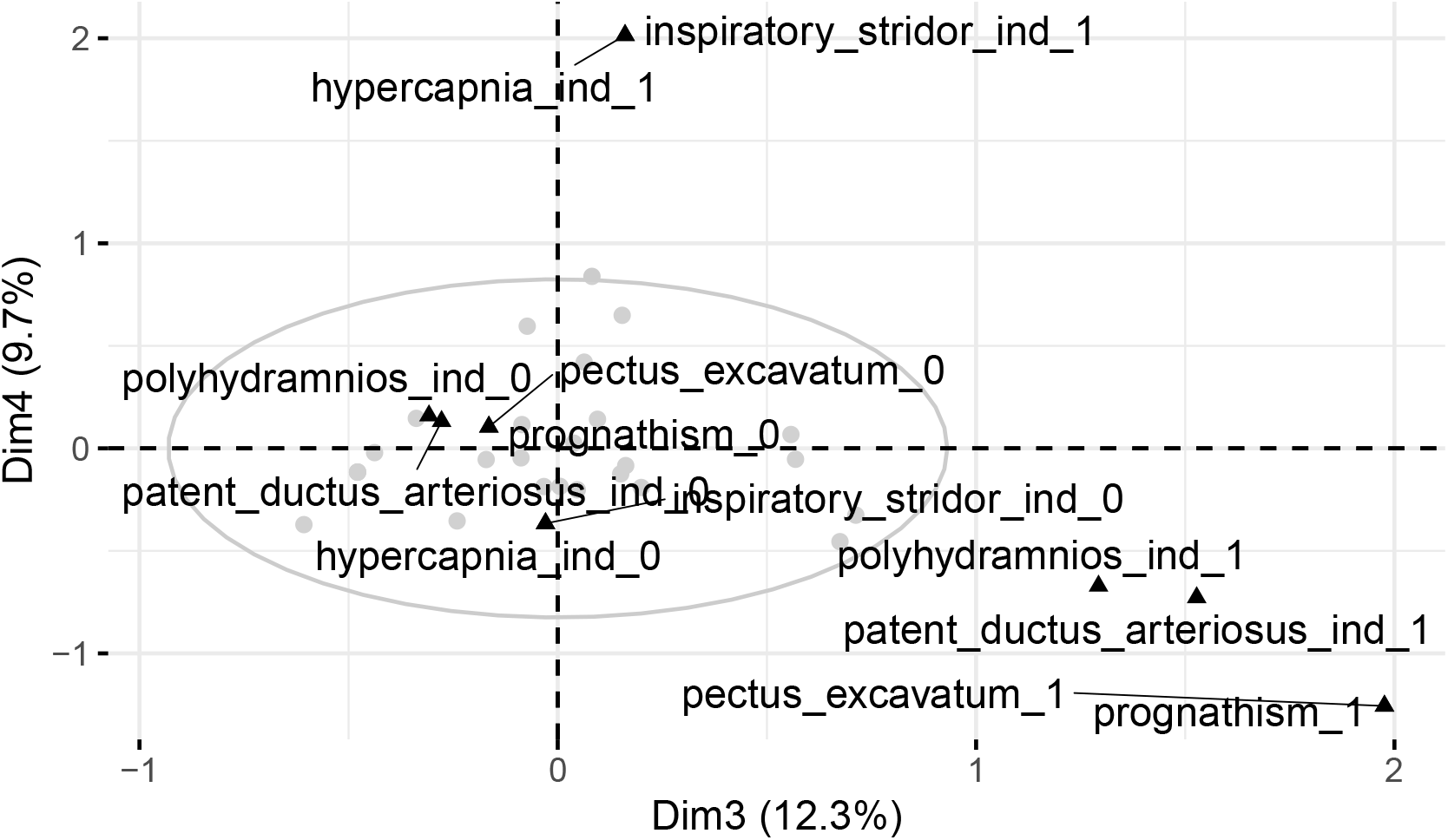
A bi-plot of principal axes 3 and 4, with grey circles indicating the individual’s projected position and black triangles indicating the variables. A 95% confidence ellipse is plotted in grey around the origin.

**Figure 5.**
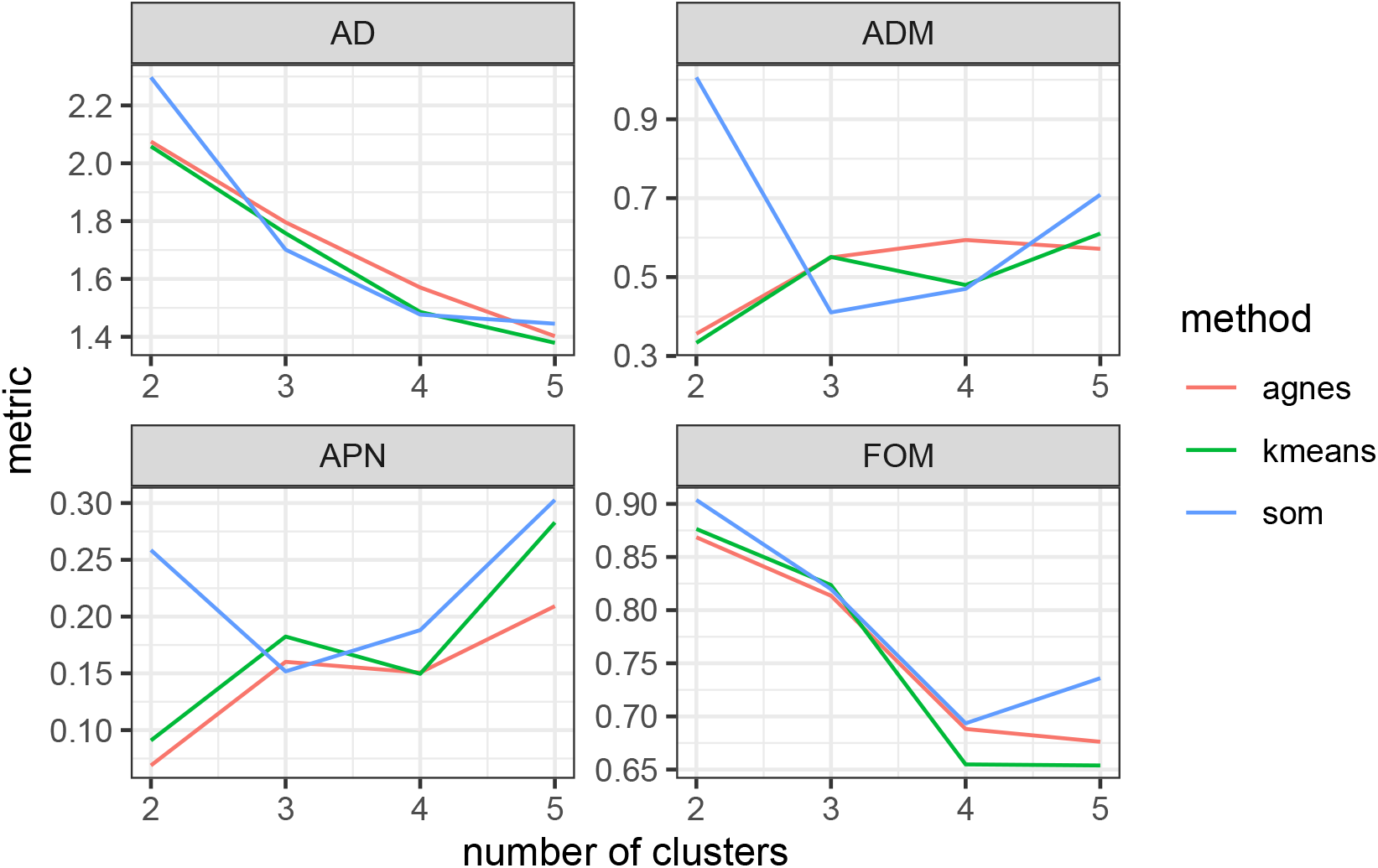
cluster validation analysis showing various metrics for cluster evaluation from 2 to 5 clusters. The most optimal number of clusters are selected by picking the lowest value from one or many of these comparable metrics.

**Figure 6.**
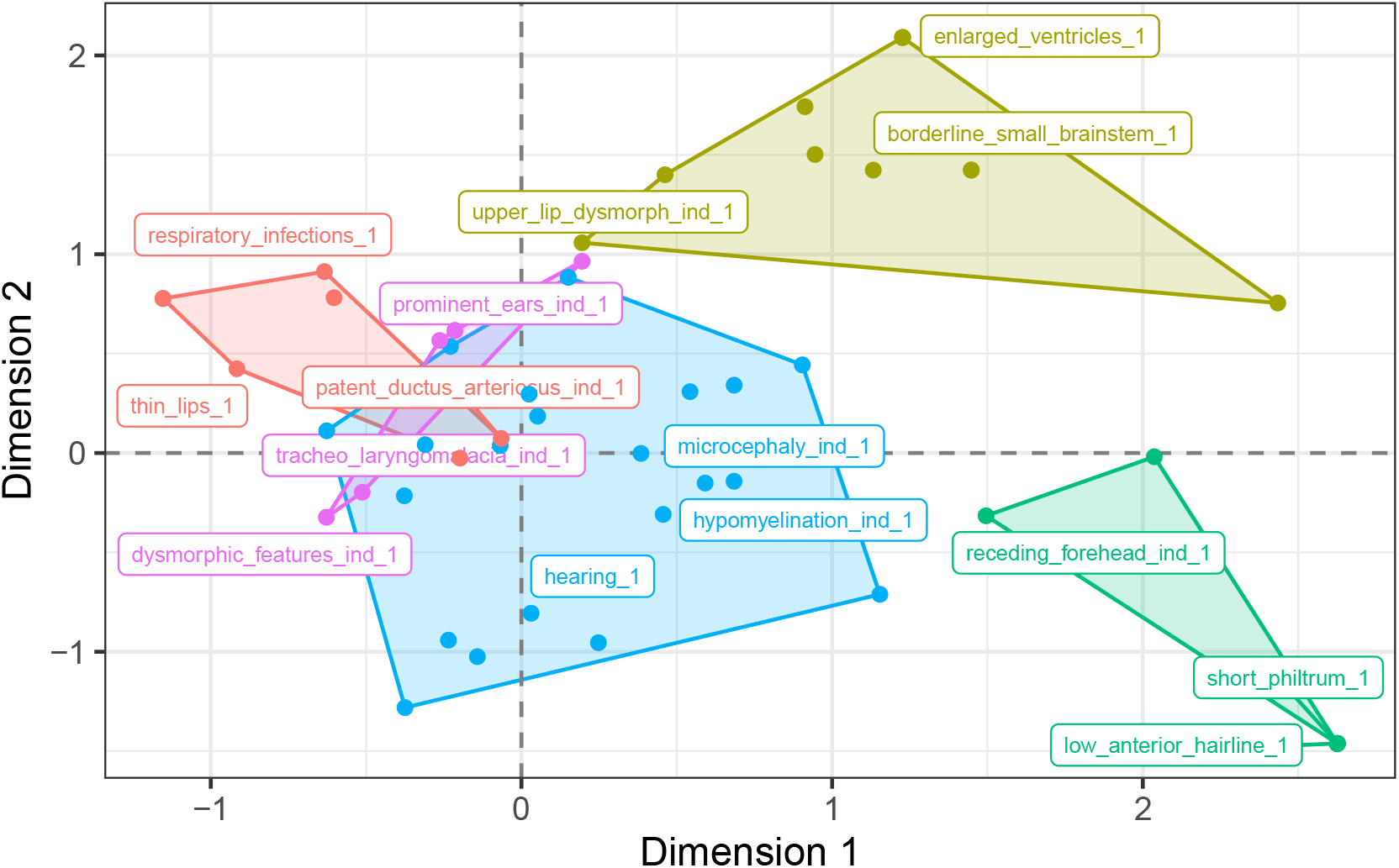
A projection of features on the first two principal components with k-means cluster assignment indicated by colour. A random sample of three labels per cluster are printed to provide a representation of each cluster’s characteristics.

### Survival Analysis

The Kaplan-Meier estimator shows a range of survival times (0, 4562 days) with median survival time calculated as 150 days. At time 0 the survival probability was 0.82 (95% CI: 0.69, 0.98). Survival probability after one year of 0.403 (95% CI: 0.252, 0.644) and survival probability at close to ten years of 0.24 (95% CI: 0.11, 0.52) See Table 10.

A comparative box plot of survival times (Figure 8) by variant type shows significant outliers for compound heterozygous *missense + splice* and homozygous *missense* variants. However, even within homozygous missense variants the survival time varies greatly. Other missense variants, such as heterozygous missense with gene deletion do not show the same increased survival time as noted by Magini et al. (2019).

**Figure 7.**
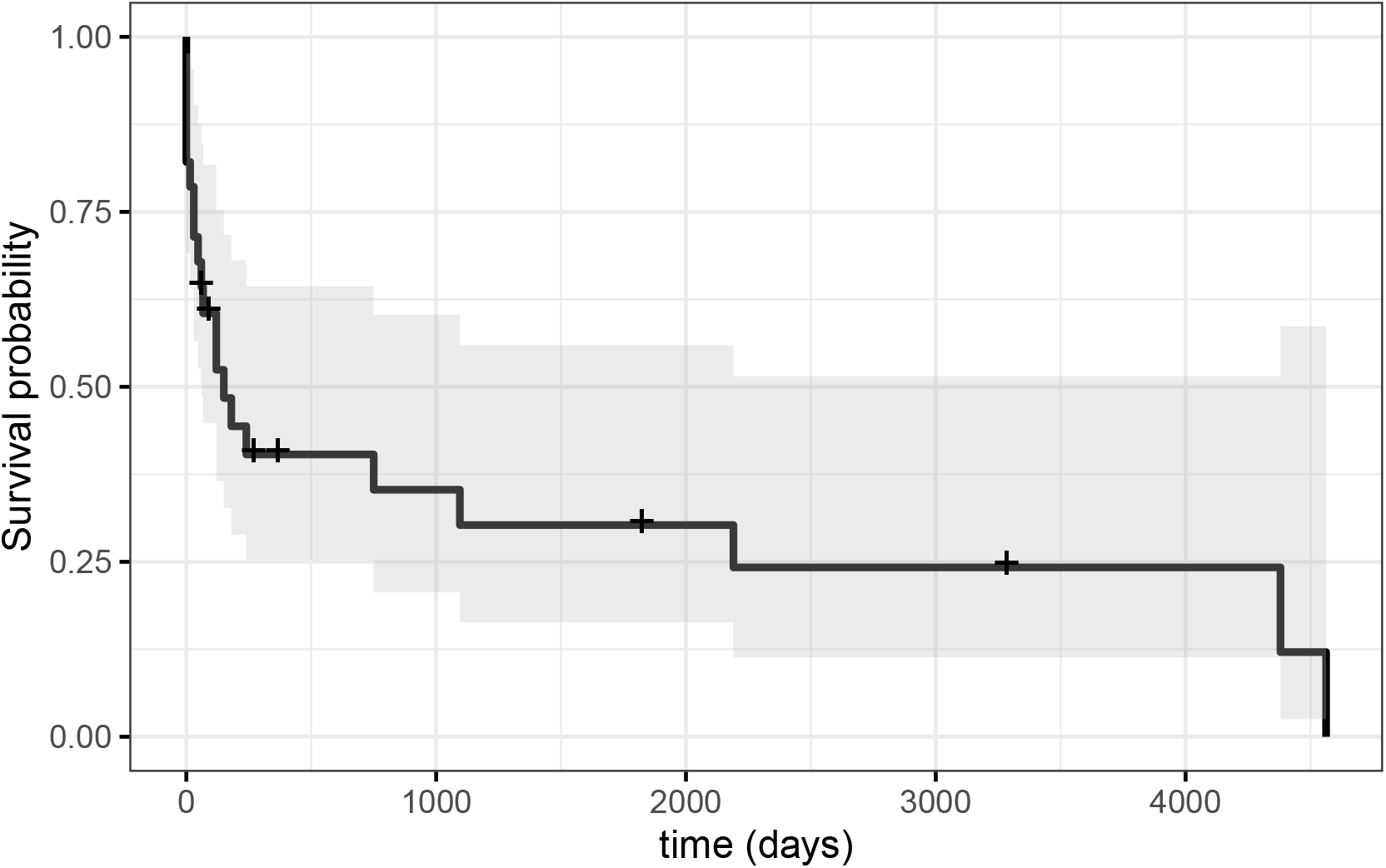
Kaplan Meier curve with the black line representing the survival probability and with grey shading indicating the 95% confidence interval. Cross icons represent right-censoring

**Figure 8.**
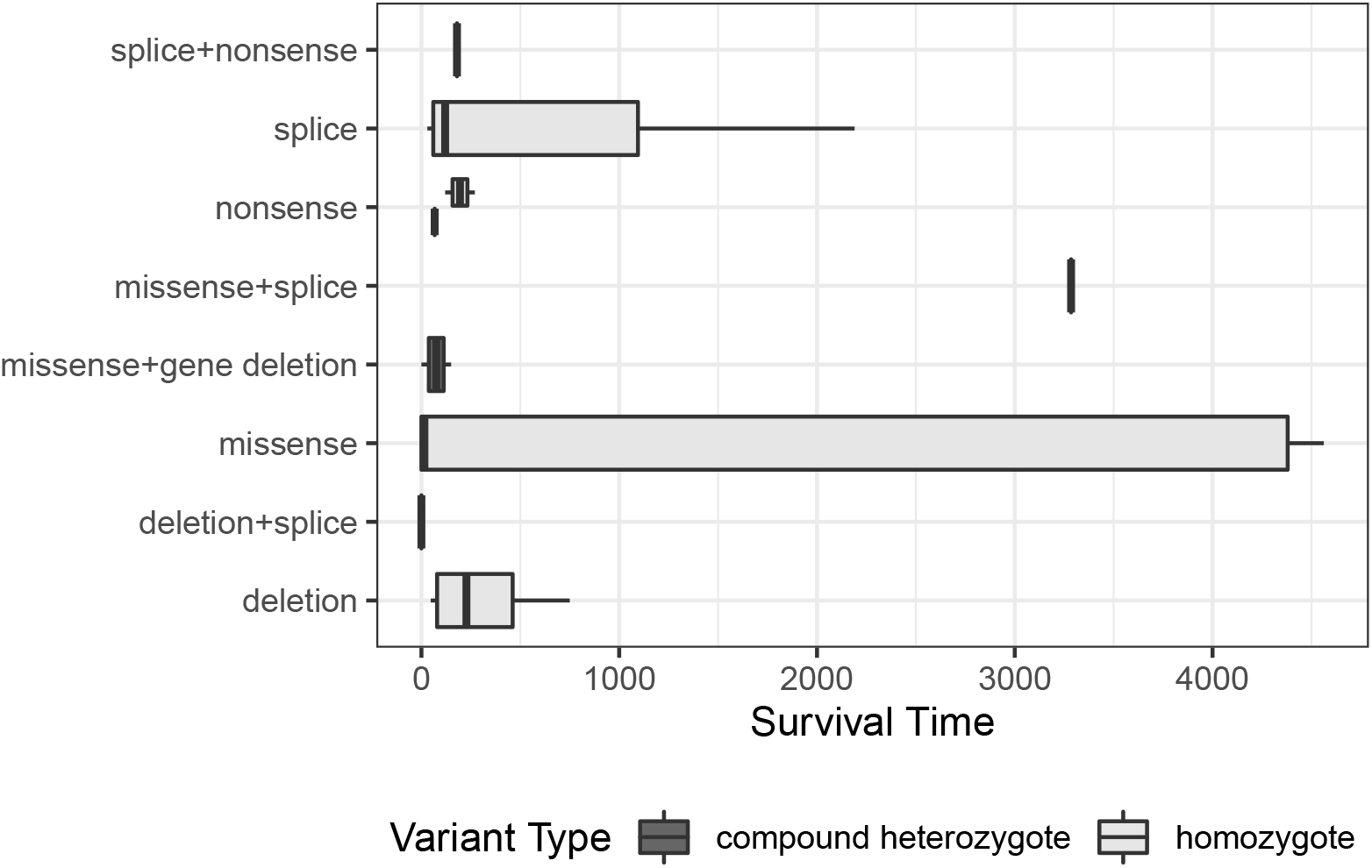
A compariative box plot showing the range of survival times for subjects split by the type of variant. Green bars indicate homozygous variants, while red indicates compound heterozygous variants.

A log-rank test was carried out to compare the survival curves of patients stratified by the various gene variants and showed a significant difference in the stratified survival curves (*χ*^2^ = 14.8, *p* = 0.04) however due to the small sample size and large number of possible variants, several had expected counts less than 5 which would create a violation in assumptions for the test.

## DISCUSSION

This meta-analysis of current research was able to consolidate data that allowed statistical analysis to be conducted on affected individuals. The use of dimension reduction techniques was shown to be successful in uncovering relevant and novel substructure in the data. This was particularly evident with the detection of the accepted typical phenotype of microcephaly, hypomyelination, arthrogryposis, simplified gyral pattern and developmental delay. After excluding two outlying remarkable cases, we identified the presence of an adjacent cluster of related features that are not well described in The Human Phenotype Ontology (Köhler et al., 2021) as shown in Table 8. It shows the prolific feature of ‘respiratory distress’ as more commonly associated with these adjacent features such as vocal cord palsy, tracheostomy, tube feeding and Gastroesophageal reflux disease (GERD). These features are present in the case reported data, but not well described in HPO and help account for the variation seen in the clinical phenotype in the first few principal dimensions.

The heterogeneous variation in the data was clearly evident with the first four principal axes being required to explain more than 50% of the variation in the data set. Visual inspection shows the primary sources of variation in the clinical phenotype are explained by a few individuals with very different clinical features. Many of these exhibit highly specific facial or anatomical dysmorphisms. While coarse facial features are a core presentation, those with a milder course had clinical reports that articulated these dysmorphisms in much more detail. Additionally, talipes appears as an isolated feature without the typical microcephaly and hypomyelination, however there was significant heterogeneity in how feet contractures were reported in clinical case reports. This is particularly evident in rocker-bottom feet which was found closely related in our analysis to core features despite the prevalence in HPO being quite low (2/21), suggesting possible under-reporting or confounded reporting of this feature. Another core identified feature is bilateral cleft lip. This condition has a reported frequency of just 2/21 (Table 8) and may be an emerging feature for a more typical presentation given its close relationship with other classic features. Features isolated to just one or few cases in our consolidated data have been labelled as ‘Isolated’ in Table 8. An optimal choice of both 2 and 5 clusters were recommended in the analysis. A partition of the data with just two clusters would likely be useful in separating outliers from other data. In this analysis a larger number (5) was selected to align the goals of the analysis which was to uncover distinct groups of features.

In terms of patient survival, the first few months of life show a sharp decline in survival probability confirming early demise in the infant period as a key factor. A small number of cases show survival into childhood and in some cases into the second decade of life. This work identifies a one year survival rate of 0.403 (95% CI: 0.252, 0.644). On the other hand, a naive estimate using just the number of deceased patients at this time as proportion of the study population is 0.43 (1 - 16/28). This highlights the importance of applying a survival analysis technique that incorporates censoring so as not to erroneously overestimate the survival probability. The current literature suggests the type of gene variant has an impact on survival time, which seems to be supported in this work. Through a comparison of multiple survival curves, stratified by variation type, the limited sample size prevented definitive conclusions with many cells having expected counts less than 5.

Despite NEDMABA exhibiting a diverse and heterogeneous clinical phenotype, our analysis was able to identify meaningful substructures in this data through the use of dimension reduction techniques (MCA) and cluster analysis. This has isolated the classic NEDMABA phenotype of microcephaly, hypomyelination, arthrogryposis, simplified gyral pattern and developmental delay. Furthermore it has isolated additional distinct groupings of features that are exhibited in patients such as vocal cord paralysis, swallowing dysfunction, tube feeding and GERD. Furthermore, we were able to quantify the expected baseline survival probabilities for children with this condition as well as quantifying the significant variation in these estimates. Future work is required with the benefit of more cases, to better understand the differences in survival curves for other factors associated with genotype.

## Data Availability

All data produced are available online at https://doi.org/10.5281/zenodo.7071092

## APPENDIX

**Table 8.** A mapping table of phenotype terms from The Human Phenotype Ontology (HPO) with clusters defined in this analysis. Not all clinical features defined in the analysis are present in the HPO terms

| HPO Data |  |  | Derived Clusters |
| --- | --- | --- | --- |
| HPO Term Name | Category | Frequency | Cluster<sup>1</sup> |
| Adducted thumb | Limbs | 1/21 | Isolated |
| Simplified gyral pattern | Nervous System | 12/18 | Core |
| Hypertonia | Musculature | 9/15 | Core |
| Seizure | Nervous System | 10/17 | Core |
| Hypotonia | Musculature | 4/15 | Core |
| Global developmental delay | Nervous System | 7/7 | Core |
| Highly arched eyebrow | Skin, Hair, and Nails | 2/21 | Isolated |
| Cerebellar hypoplasia | Nervous System | 10/21 | Core |
| Neonatal respiratory distress | Respiratory System | 14/17 | Adjacent |
| Bifid uvula | Head and neck | 1/21 | Isolated |
| Deep philtrum | Head and neck | 1/21 | Isolated |
| CNS hypomyelination | Nervous System | 7/19 | Core |
| Lumbar hypertrichosis | Skin, Hair, and Nails | 1/21 | Isolated |
| Birth length less than 3rd percentile | Growth | 6/11 | Core |
| Hypoplasia of the brainstem | Nervous System | 3/20 | Isolated |
| Tented upper lip vermilion | Head and neck | 3/21 | Isolated |
| Hypotelorism | Eye | 1/21 | Isolated |
| Short palpebral fissure | Head and neck | 4/21 | Outlying |
| Bilateral cleft lip | Head and neck | 2/21 | Core |
| Diabetes mellitus | Endocrine | 2/19 | Isolated |
| Single transverse palmar crease | Skin, Hair, and Nails | 1/21 | Isolated |
| Epicanthus | Head and neck | 5/21 | Outlying |
| Low anterior hairline | Head and neck | 2/21 | Outlying |
| Arthrogryposis multiplex congenita | Connective tissue | 17/20 | Core |
| Progressive microcephaly | Nervous System | 9/10 | Core |
| Microcephaly | Nervous System | 15/21 | Core |
| Thin upper lip vermilion | Head and neck | 2/21 | Outlying |
| Thin vermilion border | Head and neck | 3/21 | Isolated |
| Intrauterine growth retardation | Growth | 12/19 | Core |
| Posteriorly rotated ears | Ear | 2/21 | Adjacent |
| Narrow forehead | Head and neck | 1/21 | Isolated |
| Sloping forehead | Head and neck | 4/21 | Isolated |
| Smooth philtrum | Head and neck | 2/21 | Isolated |
| Short philtrum | Head and neck | 2/21 | Outlying |
| Premature birth | Prenatal and Birth | 7/18 | Core |
| Mandibular prognathia | Head and neck | 2/21 | Outlying |
| Wide intermamillary distance | Breast | 1/21 | Isolated |
| Depressed nasal bridge | Head and neck | 1/21 | Outlying |
| Downslanted palpebral fissures | Head and neck | 2/21 | Outlying |
| Short neck | Head and neck | 1/21 | Isolated |
| Bulbous nose | Head and neck | 2/21 | Isolated |
| Protruding ear | Ear | 2/21 | Outlying |
| Rocker bottom foot | Limbs | 2/21 | Core |

<sup>1</sup>Clusters defined in this analysis mapped to HPO Terms
Source: The Human Phenotype Ontology (HPO) https://hpo.jax.org/app/browse/disease/OMIM:618622
Reference: Köhler S, Gargano M, Matentzoglou N, Carmody LC, Lewis-Smith D, Vasilevsky NA, Danis D, Balagura G, Baynam G, Brower AM, Callahan TJ, Chute CG, Est JL, Galer PD, Ganesan S, Griese M, Haimel M, Pazmandi J, Hanauer M, Harris NL, Hartnett MJ, Hastreiter M, Hauck F, He Y, Jeske T, Kearney H, Kindle G, Klein C, Knoflach K, Krause R, Lagorce D, McMurry JA, Miller JA, Munoz-Torres MC, Peters RL, Rapp CK, Rath AM, Rind SA, Rosenberg AZ, Segal MM, Seidel MG, Smedley D, Talmy T, Thomas Y, Wiafe SA, Xian J, Yüksel Z, Helbig I, Mungall CJ, Haendel MA, Robinson PN. *The Human Phenotype Ontology in 2021*. Nucleic Acids Res. 2021 Jan 8;49(D1):D1207-D1217. doi: 10.1093/nar/gkaa1043. PMID: 33264411; PMCID: PMC7778952.

**Table 9.** Assignment of clinical features with their cluster. These clusters represent similar features projected onto the first four MCA principal axes. Clusters that are on opposing sides of an axes contract with each other and account for the largest amount of variation in the data.

| cluster | variable |
| --- | --- |
| 1 | polyhydramnios |
| 1 | short_palpebral_fissures |
| 1 | patent_ductus_arteriosus |
| 1 | thin_lips |
| 1 | prognathism |
| 1 | pectus_excavatum |
| 1 | respiratory_infections |
| 2 | vocal_cord_palsy |
| 2 | respiratory_distress |
| 2 | tracheostomy |
| 2 | posteriorly_angulated_ears |
| 2 | upper_lip_dysmorph |
| 2 | tube_feed |
| 2 | gerd |
| 2 | decreased_craniofacial_ratio |
| 2 | short_upturned_nose |
| 2 | borderline_small_brainstem |
| 2 | enlarged_ventricles |
| 3 | epicanthal_folds |
| 3 | receding_forehead |
| 3 | spastic_tetraparesis |
| 3 | intubation |
| 3 | low_anterior_hairline |
| 3 | short_philtrum |
| 3 | hirsutism |
| 4 | microcephaly |
| 4 | sgp |
| 4 | iugr |
| 4 | talipes |
| 4 | rocker_bottom_feet |
| 4 | contractures_hands_deform |
| 4 | arthrogryposis_contractures_other |
| 4 | cleft_lip |
| 4 | coarse_facial_features |
| 4 | hypomyelination |
| 4 | cerebellar_hypoplasia |
| 4 | small_anterior_fontanel |
| 4 | thin_corpus_collosum |
| 4 | feeding_swallow_dysfxn |
| 4 | hearing |
| 4 | seizure |
| 4 | agenesis_of_corpus_callosum |
| 4 | severe_developmental_delay |
| 4 | at_birth_sga |
| 4 | lissencephaly_and_cerebellar_hypoplasia |
| 4 | eeg_normal |
| 5 | hypercapnia |
| 5 | inspiratory_stridor |
| 5 | oligohydramnios |
| 5 | prominent_ears |
| 5 | dysmorphic_features |
| 5 | tracheo_laryngomalacia |

**Table 10.** Survival probabilities derived from Kaplan-Meier estimation for the range of recorded survival times, including 95% Confidence Interval.

| time | n.risk | n.event | n.censor | surv | std.err | upper | lower |
| --- | --- | --- | --- | --- | --- | --- | --- |
| 0.0 | 28 | 5 | 0 | 82.14% | 0.09 | 97.63% | 69.11% |
| 15.0 | 23 | 1 | 0 | 78.57% | 0.10 | 95.34% | 64.75% |
| 30.0 | 22 | 2 | 0 | 71.43% | 0.12 | 90.28% | 56.51% |
| 47.0 | 20 | 1 | 0 | 67.86% | 0.13 | 87.56% | 52.59% |
| 60.0 | 19 | 1 | 1 | 64.29% | 0.14 | 84.73% | 48.78% |
| 67.0 | 17 | 1 | 0 | 60.50% | 0.15 | 81.72% | 44.80% |
| 90.0 | 16 | 0 | 1 | 60.50% | 0.15 | 81.72% | 44.80% |
| 120.0 | 15 | 2 | 0 | 52.44% | 0.18 | 75.17% | 36.58% |
| 150.0 | 13 | 1 | 0 | 48.40% | 0.20 | 71.70% | 32.68% |
| 180.0 | 12 | 1 | 0 | 44.37% | 0.22 | 68.09% | 28.91% |
| 240.0 | 11 | 1 | 0 | 40.34% | 0.24 | 64.37% | 25.28% |
| 270.0 | 10 | 0 | 1 | 40.34% | 0.24 | 64.37% | 25.28% |
| 365.0 | 9 | 0 | 1 | 40.34% | 0.24 | 64.37% | 25.28% |
| 750.0 | 8 | 1 | 0 | 35.29% | 0.27 | 60.31% | 20.66% |
| 1095.0 | 7 | 1 | 0 | 30.25% | 0.31 | 55.97% | 16.35% |
| 1825.0 | 6 | 0 | 1 | 30.25% | 0.31 | 55.97% | 16.35% |
| 2190.0 | 5 | 1 | 0 | 24.20% | 0.39 | 51.51% | 11.37% |
| 3285.0 | 4 | 0 | 2 | 24.20% | 0.39 | 51.51% | 11.37% |
| 4380.0 | 2 | 1 | 0 | 12.10% | 0.81 | 58.65% | 2.50% |
| 4562.5 | 1 | 1 | 0 | 0.00% | *Inf* | NA | NA |

**Table 11.** Data dictionary of the final data set used for the analysis. This combines the case reports and supplied data for available and references studies. The data has been transformed into a tidy format with formatting applied.

| variable | type |
| --- | --- |
| study | character |
| id | character |
| family | character |
| individual | factor |
| deceased | logical |
| survival_time | numeric |
| variant_type | factor |
| locus_1 | numeric |
| locus_2 | numeric |
| gender | factor |
| ethnicity | character |
| consanguineous | logical |
| termination | logical |
| birth_gestation | numeric |
| route_vaginal_vs_c_sect | factor |
| birth_weight | numeric |
| birth_ofc | numeric |
| birth_length | numeric |
| age_at_demise | numeric |
| age_at_last_follow_up | numeric |
| microcephaly_ind | factor |
| sgp_ind | factor |
| iugr_ind | factor |
| talipes_ind | factor |
| rocker_bottom_feet_ind | factor |
| contractures_hands_deform_ind | factor |
| vocal_cord_palsy_ind | factor |
| arthrogryposis_contractures_other_ind | factor |
| hypercapnia_ind | factor |
| cleft_lip_ind | factor |
| inspiratory_stridor_ind | factor |
| polyhydramnios_ind | factor |
| coarse_facial_features_ind | factor |
| oligohydramnios_ind | factor |
| respiratory_distress_ind | factor |
| tracheostomy_ind | factor |
| prominent_ears_ind | factor |
| posteriorly_angulated_ears_ind | factor |
| prominent_philtrum_ind | factor |
| upper_lip_dysmorph_ind | factor |
| hypomyelination_ind | factor |
| cerebellar_hypoplasia_ind | factor |
| short_palpebral_fissures_ind | factor |
| epicanthal_folds_ind | factor |
| tube_feed_ind | factor |
| dysmorphic_features_ind | factor |
| patent_ductus_arteriosus_ind | factor |
| small_anterior_fontanel_ind | factor |
| receding_forehead_ind | factor |
| tracheo_laryngomalacia_ind | factor |
| gerd_ind | factor |
| thin_corpus_collosum_ind | factor |
| feeding_swallow_dysfxn_ind | factor |
| seizures_age_started | character |
| seizure_type_s | character |
| eeg | character |
| sleeping_problem | character |
| behavioral_issues | character |
| skeletal_changes_scoliosis | numeric |
| hip_luxation | numeric |
| development_head_control | character |
| sitting_unsupported | character |
| crawling | character |
| walking | character |
| speech | character |
| iq_int_disability | character |
| cp | character |
| performance | character |
| neuro_exam_eye_mvts_nystagmus | character |
| strabismus | character |
| feeding_swallow_dysfxn | character |
| gag | character |
| tone | character |
| strength | character |
| sensory_deficits | character |
| abnormal_mvts | character |
| dtr | character |
| other_neuro_signs | character |
| sensory_vision | character |
| hearing | character |
| other_organs | character |
| endocrinology | character |
| functional_tests | character |
| metabolic_investigation | character |
| cardiology | character |
| seizure | numeric |
| agenesis_of_corpus_callosum | factor |
| abnormal_pattern_of_cerebral_sulci | factor |
| left_sided_gallbladder | factor |
| suspected_brain_stem_dysfunction | factor |
| spastic_tetraparesis | factor |
| severe_developmental_delay | factor |
| acidosis | factor |
| small_thorax | factor |
| cyanosis | factor |
| intubation | factor |
| apnea | factor |
| mr_is_reportedly_with_migrational_defect | factor |
| at_birth_sga | factor |
| epilepsy_and_apnea | factor |
| choanal_atresia_surgeries_for_transposition_of_the_great_arteries | factor |
| interventricular_and_interatrial_defects | factor |
| smooth_philtrum | factor |
| thin_lips | factor |
| bilateral_simian_creases | factor |
| low_anterior_hairline | factor |
| short_philtrum | factor |
| prominent_nose_bridge | factor |
| depressed_nasal_bridge | factor |
| prominent_suture_ridges | factor |
| hypotelorism | factor |
| hypertrichosis_of_lower_back | factor |
| decreased_craniofacial_ratio | factor |
| there_is_overlapping_of_the_sutures | factor |
| bitemporal_narrowing | factor |
| short_upturned_nose | factor |
| short_neck | factor |
| upward_sweep_of_hair | factor |
| arched_eye_brows | factor |
| bulbous_nose | factor |
| prominent_lower_lip | factor |
| broad_chin | factor |
| prognathism | factor |
| asd | factor |
| left_multicystic_dysplastic_kidney | factor |
| anteriorly_placed_anus | factor |
| dilated_cardiomyopathy | factor |
| cryptorchidism | factor |
| hirsutism | factor |
| intestinal_malrotation | factor |
| dysphagia | factor |
| intermittent_sinus_bradycardia | factor |
| bilateral_staphylomas | factor |
| lissencephaly | factor |
| echogenic_choroid_plexus_on_head_us | factor |
| head_us_mild_ventriculomegaly | factor |
| brain_parenchyma_appears_homogeneous_without_evidence_of_focal_lesion | factor |
| hypoplastic_cerebellum | factor |
| borderline_small_brainstem | factor |
| abnormal_cerebellar_folia | factor |
| mild_vermian_hypoplasia | factor |
| asymmetry_of_lateral_ventricles | factor |
| lissencephaly_and_cerebellar_hypoplasia | factor |
| signs_of_hypoxic_encephalopathy | factor |
| mild_dilation_of_virchow_robins_spaces | factor |
| pectus_excavatum | factor |
| widely_spaced_nipples | factor |
| respiratory_infections | factor |
| enlarged_ventricles | factor |
| eeg_normal | numeric |
| bifid_uvula | factor |
| under_developed_cerebellar_inferior_vermis | factor |

